# Moderators of changes in smoking, drinking, and quitting behaviour associated with the first Covid-19 lockdown in England

**DOI:** 10.1101/2021.02.15.21251766

**Authors:** Sarah E. Jackson, Emma Beard, Colin Angus, Matt Field, Jamie Brown

## Abstract

**Aim:** To estimate changes in smoking, drinking, and quitting behaviour from before to during the first Covid-19 lockdown in England, and whether changes differed by age, sex, or social grade.

**Design:** Representative cross-sectional surveys of adults, collected monthly between August 2018 and July 2020.

**Setting:** England.

**Participants:** 36,980 adults (≥18y).

**Measurements:** Independent variables were survey month (pre-lockdown: August-February vs. lockdown months: April-July) and year (pandemic: 2019/20 vs. comparator: 2018/19). Smoking outcomes were smoking prevalence, cessation, quit attempts, quit success, and use of evidence-based or remote cessation support. Drinking outcomes were high-risk drinking prevalence, alcohol reduction attempts, and use of evidence-based or remote support. Moderators were age, sex, and occupational social grade (ABC1=more advantaged/C2DE=less advantaged).

**Findings:** Relative to changes over the same time period in 2018/19, lockdown was associated with significant increases in smoking prevalence (+24.7% in 2019/20 vs. 0.0% in 2018/19, OR_adj_=1.35[95%CI=1.12-1.63]) and quit attempts (+39.9% vs. −22.2%, OR_adj_=2.48[1.76-3.50]) among 18-34 year-olds, but not older groups. Increases in cessation (+156.4% vs. −12.5%, OR_adj_=3.08[1.86-5.09]) and the success rate of quit attempts (+99.2% vs. +0.8%, OR_adj_=2.29[1.31-3.98]) were also observed, and did not differ significantly by age, sex, or social grade. Lockdown was associated with a significant increase in high-risk drinking prevalence across all sociodemographic groups (+39.5% vs. −7.8%, OR_adj_=1.80[1.64-1.98]), with particularly high increases among women (OR_adj_=2.17[1.87-2.53]) and social grades C2DE (OR_adj_=2.34[2.00-2.74]). Alcohol reduction attempts increased significantly among high-risk drinkers from social grades ABC1 (OR_adj_=2.31[1.78-3.00]) but not C2DE (OR_adj_=1.25[0.83-1.88]), with larger increases among those aged 18-34 (OR_adj_=2.56[1.72-3.81]) and ≥60 (OR_adj_=1.43[1.05-1.95]) than 35-59 (OR_adj_=2.51[1.51-4.18]). There were few significant changes in use of support for smoking cessation or alcohol reduction, although samples were small.

**Conclusions:** In England, the first Covid-19 lockdown was associated with increased smoking prevalence among younger adults, and increased high-risk drinking prevalence among all adults. Smoking cessation activity also increased: more younger smokers made quit attempts during lockdown and more smokers quit successfully. Socioeconomic disparities in drinking behaviour were evident: high-risk drinking increased by more among women and those from less advantaged social grades (C2DE) but the rate of reduction attempts increased only among the more advantaged social grades (ABC1).

## Introduction

In order to suppress transmission of the SARS-CoV-2 virus, governments around the world have implemented guidelines and legislation to restrict social interaction, including advice to stay at home (‘lockdown’ restrictions). These restrictions may have influenced smoking, drinking, and quitting behaviours in various ways. Some people may have used tobacco or alcohol as a means of coping with increased stress or boredom. Others may have taken the opportunity to quit smoking or drink less while daily routines are disrupted and social activity is reduced. Given the different social, financial, and mental health impacts of lockdown on different sociodemographic groups (1–4), it is plausible that changes in smoking and drinking have varied according to age, sex, or socioeconomic position. Understanding how – and among which groups – smoking, drinking, and quitting behaviours have changed in response to Covid-19 lockdown restrictions is essential for building a clear picture of their public health impact and targeting messaging and support services.

The Smoking and Alcohol Toolkit Studies (5,6) are ongoing monthly cross-sectional surveys of the adult population in England that have been designed to provide insights into population-wide influences on smoking and drinking behaviour. These surveys pre-date the Covid-19 pandemic, providing the opportunity to evaluate the impact of lockdown restrictions on smoking and drinking behaviour.

Using the first monthly data collected after the first Covid-19 lockdown was implemented in England, we recently published an evaluation of population-level changes in smoking, quit attempts, drinking and alcohol reduction attempts from before (April 2019–February 2020) to during (April 2020) lockdown (7). This revealed some positive changes in smoking and drinking outcomes during lockdown: smokers and high-risk drinkers were more likely than before lockdown to report trying to quit (39.6% vs. 29.1%) or reduce their alcohol consumption (28.5% vs. 15.3%), and the rate of smoking cessation doubled. However, high-risk drinking prevalence increased (38.3% vs. 25.1%) during lockdown and there was some evidence that use of evidence-based support for alcohol reduction by high-risk drinkers decreased, with no compensatory increase in use of remote support options.

With only one month of post-lockdown data available at the time this analysis was undertaken, we were unable to establish whether these changes were sustained over time. We also lacked sufficient sample size in the post-lockdown period to explore potential moderation of these changes by key variables such as age, sex, or social grade. Understanding more about how these patterns of behaviour changed (or persisted) during the lockdown and how they differed between groups is important for informing health communications and policy decisions around provision and targeting of support for smoking cessation and alcohol reduction.

With additional data now available, this study therefore aimed to update and extend our original analysis of changes in smoking, drinking, quitting, and alcohol reduction attempts during the first Covid-19 lockdown in England (April–July 2020). We aimed to (i) establish whether the immediate changes following the outbreak of Covid-19 compared with pre-lockdown that we observed in our original analysis persisted across the entire four-month lockdown, and (ii) examine the extent to which these changes were moderated by age, sex, and social grade. Specifically, we addressed the following research questions:

1. Among adults in England, have changes in the prevalence of smoking or high-risk drinking following the outbreak of Covid-19 persisted, and if so, to what extent?
2. Among past-year smokers and after adjusting for sociodemographic characteristics and nicotine dependence, have changes in the prevalence of cessation following the outbreak of Covid-19 persisted?
3. Among past-year smokers and after adjusting for sociodemographic characteristics, have the changes in the prevalence of quit attempts following the outbreak of Covid-19 persisted?
4. Among past-year smokers attempting to quit, and after adjusting for sociodemographic characteristics and nicotine dependence, have changes in the rate of quit success or the prevalence of the use of cessation support following the outbreak of Covid-19 persisted?
5. Among high-risk drinkers and after adjusting for sociodemographic characteristics, have changes in the prevalence of alcohol reduction attempts following the outbreak of Covid-19 persisted?
6. Among high-risk drinkers attempting to reduce their alcohol consumption, and after adjusting for sociodemographic characteristics and alcohol dependence, have changes in the prevalence in the use of support for alcohol reduction following the outbreak of Covid-19 persisted?
7. Are the above changes moderated by age, sex, or occupational social grade (as an index of socioeconomic position)?

## Method

### Design

Data were drawn from the ongoing Smoking and Alcohol Toolkit Studies (5,6). The study uses a form of random location sampling to select a new sample of approximately 1,700 adults each month. Interviews are performed with one household member until quotas based on factors influencing the probability of being at home (e.g. gender, age, working status) are fulfilled. Comparisons with sales data and other national surveys show that the Toolkit studies recruit a representative sample of the population in England with regards to key demographic variables, smoking prevalence, and cigarette consumption (5,8). Data are usually collected monthly through face-to-face computer assisted interviews. However, social distancing restrictions under the Covid-19 pandemic meant that no data were collected in March 2020 and data from April 2020 onward were collected via telephone, and the lower age bound for participation was increased from 16 to 18 years due to changes in consenting procedures. The telephone-based data collection relied on the same combination of random location and quota sampling, and weighting approach as the face-to-face interviews and previous diagnostic analyses conducted on the first month of telephone data indicate good comparability between the two data collection modalities (7).

The UK Coronavirus Action Plan (9) was published on 3 March 2020, followed by government advice to practise social distancing on 16 March and behavioural restrictions enforceable by law (‘lockdown’) on 23 March. These lockdown restrictions were subsequently eased on 4 July. For the present study, we used data from respondents to the survey in the period from August 2018 through July 2020. Because the sample was restricted to people aged ≥18 years when data collection switched from face-to-face to telephone interviews (due to different consenting procedures), we excluded any participants aged 16-17 recruited before lockdown for consistency.

### Measures

#### Exposure: timing of lockdown

Our analyses focused on tests of the interaction between survey month and year, in order to establish whether any changes associated with the timing of the Covid-19 lockdown in March 2020 were over and above usual seasonal variation in our outcomes of interest. For our primary analyses, survey month was coded 0 (i.e. before lockdown) for respondents to the survey in August through February and 1 (i.e. during lockdown) for respondents to the survey in April through July. Survey year was coded 0 for respondents to the survey from August 2018 through July 2019 (i.e. comparator year) and 1 for respondents to the survey from August 2019 through July 2020 (i.e. pandemic year). The length of the pre-lockdown period was reduced from our original analysis (which included data from May rather than August (7)) to avoid overlap between the post-lockdown period in the comparator year and pre-lockdown period in the pandemic year.

Because lockdown restrictions were eased on 4 July 2020, and it was likely that much of the July data would have been obtained after this date, we conducted sensitivity analyses excluding July data from the lockdown period.

#### Outcomes

Among all adults, we assessed prevalence of current smoking and high-risk drinking (defined by an Alcohol Use Disorders Identification Test – consumption (AUDIT-C) score ≥5 (10)).

Among past-year smokers, we assessed cessation and quit attempts in the past year. Among past-year smokers who reported a quit attempt, we assessed quit success, use of evidence-based support (defined as face-to-face behavioural support, prescription medication [varenicline, bupropion, or NRT], e-cigarettes, or NRT obtained over the counter), and use of remote support (defined as telephone support, a website, or an app).

Among high-risk drinkers, we assessed alcohol reduction attempts in the past year. Among high-risk drinkers who reported a reduction attempt, we assessed use of evidence-based support (defined as face-to-face behavioural support or prescription medication) and use of remote support (defined as telephone support, a website, or an app).

See Supplementary File 1 for full details of the measures used to assess each outcome variable.

#### Covariates

Sociodemographic characteristics included age, sex, social grade, and region in England. Age was categorised as 18-24, 25-34, 35-44, 45-54, 55-64, and ≥65 years (those aged 16 or 17 who responded before April 2020 were excluded to match the age range of the sample collected during lockdown). Social grade was categorised as ABC1 (which includes managerial, professional and intermediate occupations) vs. C2DE (which includes small employers and own-account workers, lower supervisory and technical occupations, and semi-routine and routine occupations, never workers and long-term unemployed). Region in England was categorised as London, south, central, and north.

We also included measures of nicotine and alcohol dependence. Nicotine dependence was assessed with the Heaviness of Smoking Index (11), an index derived from the number of cigarettes smoked per day and time to the first cigarette of the day. Scores range from 0 (low dependence) to 6 (high dependence). Alcohol dependence was assessed with the (full, 10-item) AUDIT (10). Scores range from 0-40, with 0-7 indicating low-risk consumption, 8-19 indicating hazardous or harmful consumption, and ≥20 indicating risk of alcohol dependence (moderate-severe alcohol use disorder).

### Statistical analysis

The protocol and analysis plan was pre-registered on Open Science Framework (https://osf.io/zf6vp/). Data were analysed using SPSS v.24. Data were weighted to match the English population profile on age, social grade, region, tenure, ethnicity, and working status within sex. The dimensions are derived monthly from a combination of the English 2011 census, Office for National Statistics mid-year estimates, and an annual random probability survey conducted for the National Readership Survey. Missing cases were excluded on a per-analysis basis.

For each outcome, we analysed the prevalence by survey month (before [Aug-Feb] vs. after [April-July]) and year (pandemic [2019/20] vs. comparator [2018/19]) and constructed a logistic regression model testing the month x year interaction to test whether observed differences between months before and during lockdown were larger in the pandemic year than the comparator year. These models adjusted for time trends within years (i.e. from August=1 through July=12) and across the entire analysed period (i.e. from August 2018=1 through July 2020=24). Estimates of smoking and high-risk drinking prevalence did not have any additional adjustment, as they were weighted on important dimensions to match the population in England. Analyses of quit/reduction attempts were adjusted for age, sex, social grade, and region (to take account of small differences in the make-up of the subgroups being analysed). Analyses of smoking cessation, quit success, and use of support were adjusted for sociodemographic characteristics and level of dependence (because more dependent smokers tend to be less likely to quit and more dependent smokers/drinkers tend to be more likely to use support).

In order to test for moderation of associations, we ran a series of models for each outcome (fully adjusted for any relevant covariates, as described in the previous paragraph) in which the three-way interactions between the survey month (before vs. during lockdown), year (pandemic vs. comparator), and (i) age (18-34, 35-59, and ≥60 years), (ii) sex (male vs. female), and (iii) social grade (ABC1 vs. C2DE) were added.

Where there was evidence of moderation, we ran stratified analyses to provide more information as to the nature of the differences between groups.

To evaluate the impact of the change in modality of data collection from face-to-face (up to February 2020) to telephone (from April 2020), we replicated the diagnostic analyses we undertook in our original paper to check on the representativeness of the sample or comparability of data from wave to wave now that a large sample has been recruited via telephone. Results were very similar to those we reported in our previous paper (Supplementary File 2), and suggest it is reasonable to compare data from before and after the lockdown, despite the change in data collection method.

## Results

### Sample characteristics

A total of 36,980 adults aged ≥18 years participated in the Smoking and Alcohol Toolkit Study between August 2018 and July 2020 (mean [SD] 1,681 [30.3] per month). Sociodemographic characteristics of the sample by survey year (pandemic: 2019/20 vs. comparator: 2018/19) and month (during lockdown: April-July vs. before lockdown: August-February) are shown in Table 1.

**Table 1.** Sample characteristics by survey year and month

|  | **Comparator year (2018/19)** |  | **Pandemic year (2019/20)** |  |
| --- | --- | --- | --- | --- |
|  | August 2018-February 2019 | April-July 2019 | August 2019-February 2020<br>(before lockdown) | April-July 2020<br>(during lockdown) |
| *N* | 11836 | 6593 | 11897 | 6655 |
| Age in years, % (*n*) |  |  |  |  |
| 18-24 | 12.9 (1526) | 12.4 (817) | 12.7 (1509) | 10.9 (726) |
| 25-34 | 17.1 (2027) | 17.2 (1133) | 17.1 (2035) | 17.3 (1148) |
| 35-44 | 15.8 (1874) | 16.0 (1055) | 15.8 (1874) | 16.1 (1073) |
| 45-54 | 17.4 (2054) | 17.2 (1134) | 17.1 (2038) | 17.2 (1142) |
| 55-64 | 14.4 (1701) | 14.6 (964) | 14.6 (1739) | 15.2 (1013) |
| ≥65 | 22.4 (2654) | 22.6 (1489) | 22.7 (2702) | 23.3 (1552) |
| Sex, % (*n*) |  |  |  |  |
| Male | 49.0 (5798) | 49.3 (3250) | 49.2 (5856) | 49.3 (3279) |
| Female | 51.0 (6038) | 50.7 (3343) | 50.8 (6040) | 50.7 (3376) |
| Social grade, % (*n*) |  |  |  |  |
| ABC1 | 55.9 (6618) | 55.6 (3664) | 55.5 (6602) | 53.4 (3555) |
| C2DE | 44.1 (5218) | 44.4 (2929) | 44.5 (5295) | 44.3 (2949) |
| Missing | 0 (0) | 0 (0) | 0 (0) | 2.3 (151) |
| Region in England, % (*n*) |  |  |  |  |
| London | 15.6 (1847) | 15.7 (1036) | 15.5 (1842) | 15.4 (1023) |
| South | 26.4 (3122) | 26.3 (1731) | 26.4 (3146) | 26.6 (1769) |
| Central | 30.3 (3588) | 30.1 (1983) | 30.1 (3575) | 30.4 (2022) |
| North | 27.7 (3280) | 28.0 (1843) | 28.0 (3333) | 27.7 (1841) |

Note: All data are weighted to match the adult population in England on age, social grade, region, tenure, ethnicity, and working status within sex. In some cases, subgroup *ns* do not sum to the total *n* due to rounding.

### Associations of lockdown with changes in smoking outcomes

Table 2 shows changes in the prevalence of current smoking, cessation, quit attempts, quit success, and use of cessation support from before (August-February 2019/20) to during the Covid-19 lockdown (April-July 2020) relative to changes in these variables over the previous year (August-February 2018/19 to April-July 2019). Figure 1 shows the monthly prevalence of smoking outcomes across the entire study period. Supplementary File 3, Table 1 summarises tests of moderation of associations between the Covid-19 lockdown and changes in smoking outcomes by age, sex, and social grade.

**Table 2.** Month (August-February vs. April-July) x year (2018/19 vs. 2019/20) interactions for smoking outcomes

|  | Prevalence: % [95% CI] |  |  |  | Month x year interaction |  |  |  |
| --- | --- | --- | --- | --- | --- | --- | --- | --- |
|  | 2018/19 |  | 2019/20 |  | OR [95% CI] | *p* | OR<sub>adj</sub><br>[95% CI] | *p* |
|  | Aug-Feb | Apr-Jul | Aug-Feb | Apr-Jul |  |  |  |  |
| Smoking prevalence<sup>1</sup> | 16.9<br>[16.2-17.5] | 15.9<br>[15.1-16.8] | 16.1<br>[15.4-16.7] | 16.4<br>[15.5-17.3] | 1.09<br>[0.97-1.23] | 0.133 | 1.09<br>[0.97-1.23] | 0.129 |
| Age 18-34 | 22.4<br>[21.1-23.8] | 22.3<br>[20.4-24.1] | 21.5<br>[20.2-22.9] | 26.8<br>[24.8-28.8] | 1.35<br>[1.12-1.62] | 0.002 | 1.35<br>[1.12-1.63] | 0.002 |
| Age 35-59 | 17.5<br>[16.4-18.6] | 15.8<br>[14.4-17.1] | 16.4<br>[15.3-17.4] | 15.5<br>[14.1-16.8] | 1.06<br>[0.89-1.27] | 0.522 | 1.06<br>[0.89-1.28] | 0.513 |
| Age ≥60 | 9.9<br>[8.9-10.9] | 9.6<br>[8.2-10.9] | 10.0<br>[9.0-11.0] | 7.9<br>[6.7-9.1] | 0.81<br>[0.61-1.06] | 0.127 | 0.81<br>[0.61-1.07] | 0.129 |
| Cessation<sup>2</sup> | 4.0<br>[3.2-4.9] | 3.5<br>[2.4-4.6] | 3.9<br>[3.0-4.7] | 10.0<br>[8.2-11.8] | 3.16<br>[1.91-5.22] | <0.001 | 3.08<br>[1.86-5.09] | <0.001 |
| Quit attempts<sup>2</sup> | 28.8<br>[26.9-30.8] | 28.4<br>[25.7-31.1] | 29.5<br>[27.5-31.5] | 37.6<br>[34.8-40.3] | 1.47<br>[1.18-1.84] | 0.001 | 1.45<br>[1.16-1.81] | 0.001 |
| Age 18-34 | 32.4<br>[29.2-35.6] | 25.2<br>[21.1-29.2] | 32.1<br>[28.9-35.4] | 44.9<br>[40.8-49.0] | 2.45<br>[1.74-3.45] | <0.001 | 2.48<br>[1.76-3.50] | <0.001 |
| Age 35-59 | 27.7<br>[24.7-30.7] | 33.3<br>[28.8-37.7] | 28.6<br>[25.5-31.8] | 32.6<br>[28.3-36.9] | 0.93<br>[0.65-1.32] | 0.673 | 0.92<br>[0.65-1.32] | 0.657 |
| Age ≥60 | 22.7<br>[18.2-27.3] | 25.3<br>[18.8-31.8] | 25.9<br>[21.2-30.6] | 28.3<br>[21.6-35.0] | 0.98<br>[0.55-1.77] | 0.952 | 0.96<br>[0.53-1.75] | 0.896 |
| Quit success<sup>3</sup> | 12.5<br>[9.7-15.2] | 12.6<br>[8.7-16.4] | 12.7<br>[9.9-15.4] | 25.3<br>[21.1-29.5] | 2.31<br>[1.34-3.99] | 0.003 | 2.29<br>[1.31-3.98] | 0.003 |
| Use of evidence-based support<sup>3a</sup> | 53.1<br>[49.0-57.2] | 49.2<br>[43.4-55.1] | 53.8<br>[49.7-58.0] | 44.8<br>[39.9-49.6] | 0.81<br>[0.55-1.19] | 0.290 | 0.94<br>[0.63-1.41] | 0.757 |

**Table 2. (continued)**
|  |  |  |  |  |  |  |  |  |
| --- | --- | --- | --- | --- | --- | --- | --- | --- |
| Use of remote support<sup>3b</sup> | 0.8<br>[0.1-1.5] | 3.0<br>[1.0-5.0] | 2.4<br>[1.2-3.7] | 6.8<br>[4.4-9.3] | 0.75<br>[0.20-2.83] | 0.672 | 0.71<br>[0.19-2.73] | 0.619 |

Note: All data are weighted to match the adult population in England on age, social grade, region, tenure, ethnicity, and working status within sex.
CI, confidence interval. OR, odds ratio. OR<sub>adj</sub>, adjusted odds ratio. The OR<sub>adj</sub> for smoking prevalence adjusted for trend within year (i.e. August=1 through July=12) and trend across years (i.e. August 2018=1 through July 2020=24). OR<sub>adj</sub>s for other outcomes are additionally adjusted for age, sex, social grade, region (and, for analyses of cessation, quit success, and use of support, heaviness of smoking index).
White rows show results for the whole eligible sample; shaded rows show results from stratified analyses conducted where significant three way interaction between month, year, and the indicated variable were detected.
<sup>1</sup> Among all adults (2018/19: Aug-Feb *n*=11,820, April-July *n*=6,592; 2019/20: Aug-Feb *n*=11,892, April-July *n*=6,633).
<sup>2</sup> Among past-year smokers (2018/19: Aug-Feb *n*=2,055, April-July *n*=1,081; 2019/20: Aug-Feb *n*=1,961, April-July *n*=1,209).
<sup>3</sup> Among past-year smokers who made a quit attempt (2018/19: Aug-Feb *n*=571, April-July *n*=284; 2019/20: Aug-Feb *n*=551, April-July *n*=410).
<sup>a</sup> Prescription medication, face-to-face behavioural support, nicotine replacement therapy obtained over the counter, e-cigarettes.
<sup>b</sup> Telephone support, websites, or apps.

**Figure 1.**
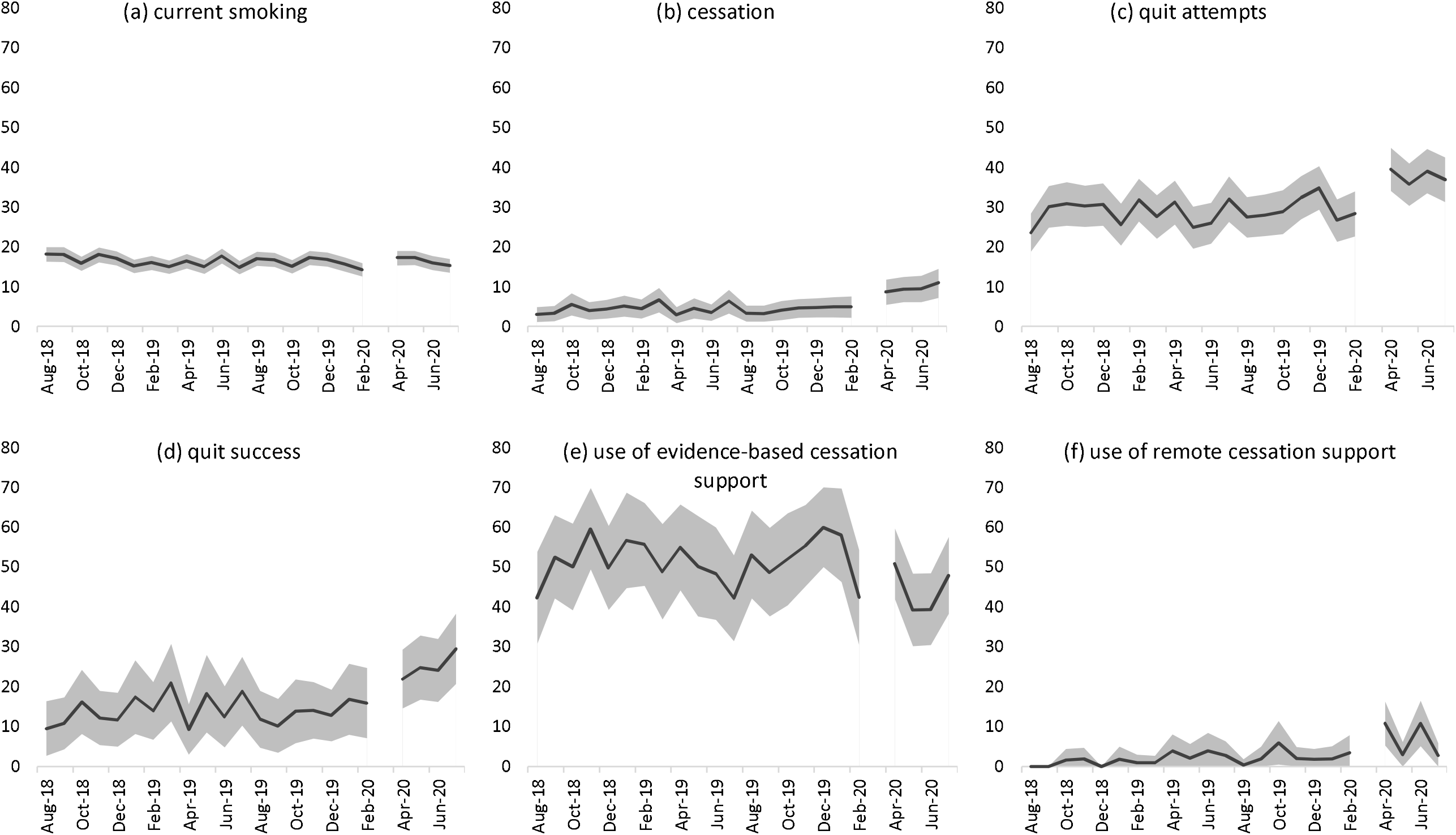
Prevalence of (a) current smoking among all adults; (b) cessation and (c) quit attempts by past-year smokers; and (d) quit success, (e) use of evidence-based cessation support and (f) use of remote cessation support by past-year smokers who made a quit attempt in England, August 2018 through July 2020. The break in the line at March 2020 indicates the timing of the Covid-19 lockdown in England (no data were collected this month). The shaded band shows the 95% confidence interval.

Among all adults, smoking prevalence was fairly stable from before to during lockdown (+0.3% from August-February 2019/20 to April-July 2020). However, a significant interaction with age (OR_adj_ 0.60 [95%CI 0.43-0.83] for ≥60 vs. 18-34) revealed an increase in smoking prevalence among 18-34 year-olds (+24.7%) that was significantly greater than changes in this group over the same time period in 2018/19 in the absence of lockdown restrictions (0.0% change).

Among past-year smokers, lockdown was associated with significant increases in the rate of cessation (+156.4%) and quit attempts (+39.9%), over and above changes over the same time period in 2018/19 (−12.5% and +0.8%, respectively). Changes in cessation did not differ significantly by smokers’ age, sex, or social grade, but the change in the prevalence of quit attempts was moderated by age (OR_adj_ 0.37 [0.23-0.61] and OR_adj_ 0.38 [0.19-0.76] for 35-59 and ≥60 vs. 18-34). Among 18-34 year-old smokers, there was a substantial increase in quit attempts (+39.9%) contrasted against a decline over the same period in 2018/19 (−22.2%). By contrast, changes among 35-59 year-olds and ≥60 year-olds from before to during lockdown were similar to those observed over the same time period in 2018/19.

Among past-year smokers who made a quit attempt, lockdown was associated with a significant increase in the success rate of quit attempts (+99.2%), which had previously been stable over the same time period in 2018/19 (+0.8%). Changes in quit success did not differ significantly by smokers’ age, sex, or social grade. There was no significant association between lockdown and use of evidence-based or remote support for smoking cessation, although the relatively small sample sizes should be noted.

### Associations of lockdown with changes in drinking outcomes

Table 3 shows changes in the prevalence of high-risk drinking, alcohol reduction attempts, and use of support for alcohol reduction from before (August-February 2019/20) to during the Covid-19 lockdown (April-July 2020) relative to changes in these variables over the previous year (August-February 2018/19 to April-July 2019). Figure 2 shows the monthly prevalence of drinking outcomes across the entire study period. Supplementary File 3, Table 2 summarises tests of moderation of associations between the Covid-19 lockdown and changes in drinking outcomes by age, sex, and social grade.

**Table 3.** Month (August-February vs. April-July) x year (2018/19 vs. 2019/20) interactions for drinking outcomes

|  | Prevalence: % [95% CI] |  |  |  | Month x year interaction |  |  |  |
| --- | --- | --- | --- | --- | --- | --- | --- | --- |
|  | 2018/19 |  | 2019/20 |  | OR [95% CI] | *p* | OR<sub>adj</sub><br>[95% CI] | *p* |
|  | Aug-Feb | Apr-Jul | Aug-Feb | Apr-Jul |  |  |  |  |
| High-risk drinking prevalence<sup>1</sup> | 26.8<br>[26.0-27.6] | 24.7<br>[23.7-25.8] | 25.8<br>[25.0-26.6] | 36.0<br>[34.8-37.1] | 1.80<br>[1.64-1.98] | <0.001 | 1.80<br>[1.64-1.98] | <0.001 |
| Male | 34.6<br>[33.4-35.8] | 33.4<br>[31.8-35.0] | 34.2<br>[33.0-35.4] | 44.7<br>[43.0-46.4] | 1.64<br>[1.45-1.86] | <0.001 | 1.64<br>[1.45-1.87] | <0.001 |
| Female | 19.4<br>[18.4-20.4] | 16.4<br>[15.1-17.6] | 17.7<br>[16.7-18.7] | 27.5<br>[26.0-29.0] | 2.17<br>[1.87-2.53] | <0.001 | 2.17<br>[1.87-2.53] | <0.001 |
| Social grade ABC1 | 31.5<br>[30.4-32.6] | 30.1<br>[28.7-31.6] | 31.2<br>[30.1-32.4] | 40.3<br>[38.7-41.9] | 1.58<br>[1.40-1.79] | <0.001 | 1.58<br>[1.40-1.79] | <0.001 |
| Social grade C2DE | 20.9<br>[19.8-22.0] | 18.0<br>[16.6-19.4] | 19.1<br>[18.0-20.1] | 31.3<br>[29.6-33.0] | 2.34<br>[2.00-2.73] | <0.001 | 2.34<br>[2.00-2.74] | <0.001 |
| Alcohol reduction attempts<sup>2</sup> | 14.8<br>[13.5-16.0] | 15.7<br>[13.9-17.5] | 15.1<br>[13.8-16.4] | 26.5<br>[24.7-28.3] | 1.88<br>[1.51-2.33] | <0.001 | 1.95<br>[1.57-2.43] | <0.001 |
| Age 18-34 | 12.5<br>[10.6-14.4] | 12.9<br>[10.1-15.7] | 11.7<br>[9.8-13.7] | 25.8<br>[22.5-29.1] | 2.52<br>[1.70-3.74] | <0.001 | 2.56<br>[1.72-3.81] | <0.001 |
| Age 35-59 | 16.0<br>[14.0-17.9] | 18.7<br>[15.8-21.6] | 18.8<br>[16.7-20.9] | 28.2<br>[25.4-30.9] | 1.40<br>[1.03-1.91] | 0.030 | 1.43<br>[1.05-1.95] | 0.023 |
| Age ≥60 | 16.5<br>[13.5-19.5] | 14.0<br>[10.1-18.0] | 13.1<br>[10.4-15.8] | 23.8<br>[20.0-27.6] | 2.51<br>[1.52-4.15] | <0.001 | 2.51<br>[1.51-4.18] | <0.001 |
| Social grade ABC1 | 17.4<br>[15.8-19.1] | 15.8<br>[13.6-18.0] | 16.8<br>[15.2-18.5] | 29.7<br>[27.3-32.1] | 2.35<br>[1.81-3.05] | <0.001 | 2.31<br>[1.78-3.00] | <0.001 |
| Social grade C2DE | 9.7<br>[7.9-11.5] | 15.6<br>[12.4-18.8] | 11.7<br>[9.6-13.7] | 21.4<br>[18.6-24.1] | 1.20<br>[0.80-1.79] | 0.389 | 1.25<br>[0.83-1.88] | 0.289 |
| Use of evidence-based support<sup>3a</sup> | 4.3<br>[2.4-6.1] | 5.9<br>[3.0-8.9] | 3.0<br>[1.4-4.6] | 1.2<br>[0.3-2.1] | 0.28<br>[0.09-0.88] | 0.030 | 0.39<br>[0.11-1.45] | 0.160 |

**Table 3. (continued)**
|  |  |  |  |  |  |  |  |  |
| --- | --- | --- | --- | --- | --- | --- | --- | --- |
| Use of remote support<sup>3b</sup> | 3.8<br>[2.1-5.6] | 5.9<br>[2.9-8.8] | 5.9<br>[3.7-8.1] | 8.0<br>[5.8-10.2] | 0.89<br>[0.37-2.11] | 0.782 | 1.08<br>[0.94-1.24] | 0.292 |
| Male | 4.1<br>[1.8-6.4] | 8.7<br>[4.2-13.2] | 5.3<br>[2.6-7.9] | 6.0<br>[3.4-8.5] | 0.51<br>[0.18-1.49] | 0.218 | 0.50<br>[0.17-1.54] | 0.229 |
| Female | 3.4<br>[0.7-6.2] | 1.1<br>[0.0-3.2] | 6.9<br>[3.1-10.8] | 10.7<br>[6.8-14.5] | 5.19<br>[0.54-49.54] | 0.153 | 13.27<br>[1.22-144.75] | 0.034 |
| Social grade ABC1 | 4.3<br>[2.2-6.4] | 4.0<br>[1.0-7.0] | 4.9<br>[2.6-7.2] | 8.8<br>[6.0-11.6] | 2.02<br>[0.67-6.08] | 0.213 | 2.38<br>[0.75-7.54] | 0.141 |
| Social grade C2DE | 2.2<br>[0.0-5.1] | 9.9<br>[3.1-16.6] | 8.9<br>[3.6-14.2] | 6.2<br>[2.7-9.8] | 0.14<br>[0.02-0.80] | 0.028 | 0.11<br>[0.02-0.82] | 0.031 |

Note: All data are weighted to match the adult population in England on age, social grade, region, tenure, ethnicity, and working status within sex.
CI, confidence interval. OR, odds ratio. OR<sub>adj</sub>, odds ratio. The OR<sub>adj</sub> for high-risk drinking prevalence is adjusted for trend within year (i.e. August=1 through July=12) and trend across years (i.e. August 2018=1 through July 2020=24). OR<sub>adj</sub>s for other outcomes are additionally adjusted for age, sex, social grade, region (and, for analyses of use of support, full AUDIT score as an indicator of dependence).
White rows show results for the whole eligible sample; shaded rows show results from stratified analyses conducted where significant three way interaction between month, year, and the indicated variable were detected.
<sup>1</sup> Among all adults (2018/19: Aug-Feb *n*=11,793, April-July *n*=6,562; 2019/20: Aug-Feb *n*=11,828, April-July *n*=6,526).
<sup>2</sup> Among high-risk drinkers (2018/19: Aug-Feb *n*=3,091, April-July *n*=1,571; 2019/20: Aug-Feb *n*=2,986, April-July *n*=2,217).
<sup>3</sup> Among high-risk drinkers who made a reduction attempt (2018/19: Aug-Feb *n*=456, April-July *n*=247; 2019/20: Aug-Feb *n*=449, April-July *n*=581).
<sup>a</sup> Prescription medication or face-to-face behavioural support.
<sup>b</sup> Telephone support, websites, or apps.

**Figure 2.**
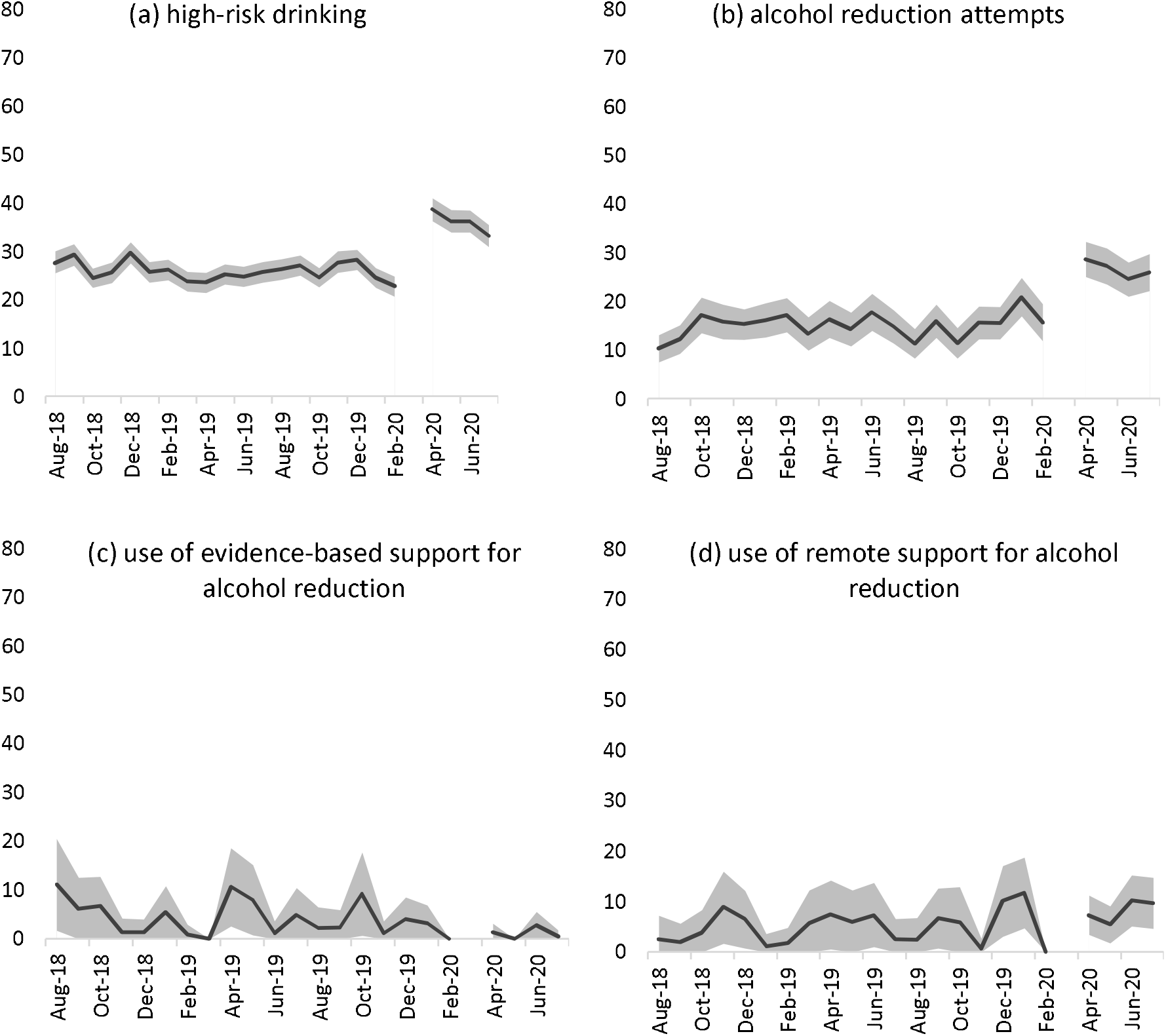
Prevalence of (a) high-risk drinking among all adults; (b) reduction attempts by high-risk drinkers; and (c) use of evidence-based support and (d) use of remote support for alcohol reduction by high-risk drinkers who made a reduction attempt in England, August 2019 through July 2020. The break in the line at March 2020 indicates the timing of the Covid-19 lockdown in England (no data were collected this month). The shaded band shows the 95% confidence interval.

Among all adults, the Covid-19 lockdown was associated with an increase in the prevalence of high-risk drinking (+39.5%) contrasted against a small decline (−7.8%) over the same time period in 2018/19. This result did not differ significantly by age, but was moderated by sex (OR_adj_ 1.32 [1.09-1.61]) and social grade (OR_adj_ 1.48 [1.21-1.80]). While increases in high-risk drinking were observed across all groups compared with small declines in 2018/19, there were greater increases among women (+55.4%) than men (+30.7%), and people from social grades C2DE (+63.9%) than ABC1 (+29.2%).

Among high-risk drinkers, lockdown was associated with a significant increase in alcohol reduction attempts (+75.5%) relative to the same time period in 2018/19 (−7.8%). This increase was moderated by age (OR_adj_ 0.55 [0.33-0.91] for 35-59 vs. 18-34) and social grade (OR_adj_ 0.54 [0.33-0.87]). Significant increases were seen across all age groups, but changes relative to the same time period in 2018/19 were smaller among those aged 35-59 (who were most likely to report alcohol reduction attempts before lockdown; +50.0%) than among those who were younger (+120.5%) or older (+81.7%). While substantial increases in the prevalence of alcohol reduction attempts were observed both in social grades ABC1 (+76.8%) and C2DE (+82.9%), alcohol reduction attempts had been fairly stable over the same time period in 2018/19 in social grades ABC1 (−9.2%) but had increased by a similar magnitude in social grades C2DE (+60.8%). Thus, the lockdown appeared only to be significantly associated with increased alcohol reduction attempts among high-risk drinkers from social grades ABC1.

Among high-risk drinkers who made an alcohol reduction attempt, lockdown was associated with a decline in use of evidence-based support (−60.0%) compared with an increase (+37.2%) over the same time period in 2018/19. This change was significant in the unadjusted model but was attenuated and no longer significant after adjustment for covariates (possibly due to the small sample sizes; see Table 3 footnote). There was no significant association overall between lockdown and use of remote support by high-risk drinkers, but there was evidence of moderation by sex (OR_adj_ 16.88 [1.30-218.96]) and social grade (OR_adj_ 0.07 [0.01-0.62]). Sample sizes were small, but significant associations were observed between lockdown and increased use of remote support among women (+55.1%, compared to a decline [-67.6%] in 2018/19) and decreased use of remote support among high-risk drinkers from social grades C2DE (−30.3%, compared to an increase [+350.0%] in 2018/19).

### Sensitivity analyses

Results of sensitivity analyses excluding data from the month of July are shown in Supplementary File 4. While effect sizes differed slightly, there were no notable differences in the pattern of results.

## Discussion

The first Covid-19 lockdown was associated with significant changes in smoking, drinking, and quitting behaviour among adults in England compared with changes across the same period a year previous. Smoking prevalence and quit attempts increased comparatively among 18-34 year-olds, but not older groups. Cessation and the success rate of quit attempts also increased comparatively, with no evidence of moderation by age, sex, or social grade. High-risk drinking prevalence increased comparatively across all groups, but particularly pronounced rises were seen in women and people from less advantaged social grades (C2DE). Alcohol reduction attempts increased significantly comparatively among high-risk drinkers from social grades ABC1 but not C2DE for whom there had been a similar increase in equivalent period a year previous, and there were smaller increases observed among 35-59 year-olds compared with 18-34 and ≥60 year-olds. There was little evidence of significant changes in use of support for smoking cessation or alcohol reduction, although sample sizes were small – particularly when the sample was stratified by age, sex, or social grade.

These results build on and extend our previous analysis of data from the first month of lockdown (7) by covering the full duration of the first lockdown in England (April-July) and exploring the extent to which changes in smoking and drinking outcomes differed by age, sex, and social grade. There are two key findings. The first is that the changes in smoking, drinking, and quitting we documented in the first month of lockdown appear to have broadly persisted throughout the four-month lockdown. The only exception was changes in use of support, which fluctuated and were not statistically significant. This suggests increases in high-risk drinking, efforts to reduce alcohol consumption and quit smoking, and success in the latter were not short-lived, acute reactions to lockdown. Figures 1 and 2 provide an indication of trends during the lockdown period and are not suggestive of any substantial decay in our observed effects over the four-month lockdown, with the possible exception of high-risk drinking prevalence. Further analyses of longer-term trends beyond the first lockdown and during subsequent periods of differing Covid-19 restrictions will provide interesting insight into the duration of these changes and the extent to which they recurred during later lockdowns.

The second key finding is that changes in smoking, drinking, and quitting behaviours have not occurred equally across all sociodemographic groups. An increase in smoking prevalence during lockdown was only evident among younger adults (age 18-34 years), with rates relatively stable among older age groups. This might be explained by differential impacts of the pandemic on younger versus older adults: several studies have shown that younger adults report higher levels of pandemic-related stress, say their lives have changed more due to the pandemic, feel more socially isolated, and have lower levels of psychological wellbeing (1,12). Many people mistakenly believe smoking relieves stress (13,14), so those experiencing lockdown-induced stress may have taken up or relapsed to smoking in an effort to cope. Given older people report being more worried about becoming seriously ill from Covid-19 (15,16), health concerns might have served as a greater deterrent from smoking at older than younger ages. The increase in quit attempts we observed was also concentrated in the younger age group. The apparent discordance between increased prevalence and increased quit attempts among younger adults does not have an obvious explanation and warrants further investigation. It may relate to substantial and unprecedented changes in demography: more than a million people were estimated to have left England during the pandemic (17). If smoking prevalence among the group leaving was lower than the population remaining, then national prevalence estimates could appear to increase despite quitting activity. The pandemic may also have negatively impacted uptake and late relapse (>1 year), which could lead to increases in prevalence despite increases in short-term (<1 year) quitting activity observed in the current study.

With regard to drinking outcomes, an increase in high-risk drinking prevalence was observed across all sociodemographic groups, but the change was greater among women than men, and among adults from social grades C2DE (less advantaged) than ABC1 (more advantaged). In addition, a significant increase in alcohol reduction attempts was only observed among high-risk drinkers from social grades ABC1: while absolute changes from before to during lockdown were similar across social grades, the change in social grades C2DE was comparable with changes over the same time period in the previous year. The greater increase in high-risk drinking among women than men has been documented in other surveys (18–20) and may reflect stress associated with an exacerbation of gender inequalities: during the pandemic, women have experienced higher rates of job loss and taken on a disproportionately greater share of housework, childcare and home schooling responsibilities (2,3,21,22). The greater increase in high-risk drinking among less socioeconomically advantaged groups has not consistently been observed, with a previous survey finding that low income was associated with drinking less than usual during the first few weeks of the lockdown, and high income and post-16 qualifications were associated with drinking more (18). Differences between these results could be due to differences in methodology (i.e. measures used to assess drinking) or timing: the present results cover the full duration of the lockdown and may reflect cumulative effects of the pandemic on less advantaged social grades. The pandemic has worsened socioeconomic inequalities (2,4,23), which may have driven the greater increase in high-risk drinking among people from less advantaged social grades and made attempting to reduce alcohol consumption less of a priority.

The present findings have implications for public health. While lockdown restrictions have been necessary to control Covid-19 transmission, they may have adversely affected population health through increased prevalence of high-risk drinking and increased uptake of or relapse to smoking among younger adults. With greater increases in high-risk drinking among adults from social grades C2DE than ABC1 and increased alcohol reduction attempts among social grades ABC1 but not C2DE, socioeconomic inequalities in health may worsen as a result of lockdown-associated drinking. We note that although increases in high-risk drinking were comparatively smaller among men and social grades ABC1 than women and social grades C2DE respectively, the former groups had higher prevalence of high-risk drinking both before and during lockdown. It will be important to monitor the extent to which changes in smoking and drinking during lockdown are sustained over the medium and long-term in order to evaluate the full public health impact of the pandemic and help to tailor future harm reduction interventions.

Strengths of this study include the large, representative sample and the broad range of data captured on smoking and drinking behaviour. The repeat cross-sectional design with data pre-dating the pandemic was also a strength, as was the comparison of changes from before to during lockdown with data from the previous year, which allowed us to rule out seasonal explanations for changes in our outcomes of interest. There were also several limitations. First, despite the large overall sample, analyses for some of the outcomes (e.g. use of support) were limited to relatively small numbers of participants (i.e. smokers/high-risk drinkers who had made a quit/reduction attempt). This resulted in estimates with wide confidence intervals, and limited statistical power to detect significant interactions with age, sex, or social grade. As such, we emphasise the need to interpret results as providing no evidence of differences between these groups, rather than evidence of no differences. Secondly, it is possible that the change in modality of data collection from face-to-face (before lockdown) to telephone interviews (during lockdown) may have contributed to some of the changes in smoking and drinking behaviour we observed. However, the diagnostic analyses we undertook comparing the face-to-face and telephone data, combined with previous studies showing a high degree of comparability between face-to-face and telephone interviews (24,25), suggest it is reasonable to compare data collected via the two methods. Finally, we did not model changes within the lockdown period, so our analyses cannot conclusively tell us whether immediate changes after lockdown was implemented were sustained, decayed, or even increased over the four months of lockdown. We plan to conduct more sophisticated interrupted time series modelling when sufficient post-lockdown data points are available. However, we provide descriptive data on monthly changes in each outcome in Figures 1 and 2 to supplement our primary analyses of aggregated data.

In conclusion, the first Covid-19 lockdown in England in March-July 2020 was associated with increased smoking prevalence among younger adults, and increased prevalence of high-risk drinking among all sociodemographic groups. Smoking cessation activity also increased: more younger smokers made quit attempts during lockdown and more smokers quit successfully. However, socioeconomic disparities in patterns of drinking behaviour were evident: high-risk drinking increased by more among women and those from less advantaged social grades but the rate of alcohol reduction attempts increased only among the more advantaged social grades.

## Data Availability

Data are available on request.

## Declarations

## Ethics approval and consent to participate

Ethical approval for the STS/ATS was granted originally by the UCL Ethics Committee (ID 0498/001). The data are not collected by UCL and are anonymised when received by UCL.

