## Supplementary File 1 for "Moderators of changes in smoking, drinking, and quitting behaviour associated with the first Covid-19 lockdown in England"

Full details of measures used to assess smoking and drinking outcomes

### Smoking outcomes

###### Smoking status

Smoking status was assessed with the question: “Which of the following best applies to you? (*a*) I smoke cigarettes (including hand-rolled) every day; (*b*) I smoke cigarettes (including hand-rolled), but not every day; (*c*) I do not smoke cigarettes at all, but I do smoke tobacco of some kind (e.g., pipe, cigar or shisha); (*d*) I have stopped smoking completely in the last year; (*e*) I stopped smoking completely more than a year ago; (*f*) I have never been a smoker (i.e. smoked for a year or more).” Current smoking was coded 1 for those who reported smoking any type of tobacco (i.e. responses *a-c*) and 0 for those who reported being a former or never smoker (responses *d-f*). Past-year smoking as coded 1 for those who reported current smoking or having stopped in the past year (responses *a-d*) and 0 for those who reported stopping more than a year ago or never smoking (responses *e-f*).

###### Smoking cessation

Among past-year smokers, cessation was coded 1 for those who reported having stopped smoking completely in the last year (response d to the measure of smoking status described above) and 0 for those who reported being a current smoker (responses a-c).

###### Attempts to stop smoking and quit success

Among past-year smokers, attempts to stop smoking were assessed with the question: “How many serious attempts to stop smoking have you made in the last 12 months? By serious attempt I mean you decided that you would try to make sure you never smoked again. Please include any attempt that you are currently making and please include any successful attempt made within the last year.” Those who reported making at least one serious quit attempt in the past year were coded 1, else they were coded 0.

Among past-year smokers who reported a quit attempt, quit success was coded 1 for those who reported having stopped smoking completely in the last year (response *d* to the measure of smoking status described above) and 0 for those who reported being a current smoker (responses *a-c*).

###### Use of support for smoking cessation

Among past-year smokers who reported making at least one quit attempt in the past year, use of cessation support in the most recent quit attempt as assessed with the question: “Which, if any, of the following did you try to help you stop smoking during the most recent serious quit attempt?” We analysed two variables: use of evidence-based support and use of remote support. Use of evidence-based support was coded 1 for those who reported using any of face-to-face behavioural support, prescription medication (varenicline, bupropion, or NRT), e-cigarettes, or NRT obtained over the counter, and 0 for those who did not report using any of these. Use of remote support was coded 1 for those who reported using telephone support, a website, or an app, and 0 for those who did not report using any of these.

### Drinking outcomes

###### High-risk drinking

Participants completed the three consumption questions of the Alcohol Use Disorders Identification Test (AUDIT-C) (1), a screening tool developed by the World Health Organization which asks about alcohol use over the past six months. The AUDIT-C classifies people scoring ≥5 as high-risk drinkers, and has demonstrated responsiveness to change, validity, high internal consistency, and good test-retest reliability across gender, age, and cultures (2–7).

###### Attempts to restrict alcohol consumption

Among high-risk drinkers, attempts to reduce alcohol consumption were assessed with the question: “How many serious attempts to cut down on your drinking alcohol have you made in the last 12 months? By serious attempt I mean you decided that you would try to make sure you reduced the amount you drank permanently. Please include any attempt that you are currently making and please include any successful attempt made within the last 12 months.” Those who reported making at least one serious reduction attempt in the past year were coded 1, else they were coded 0.

###### Use of support for alcohol reduction

Among high-risk drinkers who reported making at least one alcohol reduction attempt in the past year, use of support in the most recent attempt was assessed with the question: “Which, if any, of the following did you try to help you cut down during the most recent serious attempt?” We analysed two variables: use of evidence-based support and use of remote support. Use of evidence-based support was coded 1 for those who reported using any of face-to-face behavioural support or prescription medication (e.g. acamprosate, disulfiram, nalmefene), and 0 for those who did not report using any of these. Use of remote support was coded 1 for those who reported using telephone support, a website, or an app, and 0 for those who did not report using any of these.
