## Supplementary File 2 for "Moderators of changes in smoking, drinking, and quitting behaviour associated with the first Covid-19 lockdown in England"

Diagnostic analysis evaluating the potential impact of the change in modality of data collection from face-to-face (before the first Covid-19 lockdown) to telephone (during and after the lockdown) on the representativeness of the sample or comparability of data from wave to wave

(ii) Are there notable differences in the type of participants recruited by modality?

Our first check compared the unweighted characteristics of the samples recruited face-to-face (before the pandemic) and by telephone (after the pandemic). There were some significant differences in the characteristics of the two samples (Table 1). Relative to face-to-face sampling, the sample recruited by telephone underrepresented the younger age groups and men, and overrepresented people living in the South (outside of London). There was no significant difference by social grade.

(iii) Is more weighting required to achieve a representative sample from telephone interviews?

Our second check compared the extent of weighting applied to face-to-face and telephone samples required to achieve representation of the population in England. Weights applied to the sample recruited face-to-face before the pandemic ranged from 0.16 to 4.31, with a standard deviation of 0.41, and those applied to the sample recruited via telephone after the pandemic ranged from 0.30 to 3.71, with a standard deviation of 0.39. The narrower range and smaller standard deviation of weights applied to the telephone sample do not suggest this sample was less representative of the population in England.

(iv) Are expected associations observed?

Our final check tested whether known associations between smoking, high-risk drinking, and sociodemographic characteristics were observed when participants were recruited via telephone. The results (Table 2) confirmed that odds of high-risk drinking were higher among smokers than non-smokers and men than women, and odds of smoking were higher among those aged 18-34 than ≥35 years and among those from social grades C2DE (more disadvantaged) than ABC1 (more advantaged).

| **Table 1.** Unweighted characteristics of the samples recruited before and during the first Covid-19 lockdown in England | | | | |
| --- | --- | --- | --- | --- |
|  | | **Before lockdown (face-to-face)** | **During lockdown (telephone)** | ***p*^1^** |
| *N* | | 30289 | 6653 | - |
| Age in years, % (*n*) | |  |  |  |
|  | 18-24 | 14.0 (4251) | 9.0 (600) | <0.001 |
|  | 25-34 | 14.7 (4449) | 12.8 (851) | - |
|  | 35-44 | 14.3 (4319) | 13.2 (877) | - |
|  | 45-54 | 14.3 (4344) | 17.7 (1178) | - |
|  | 55-64 | 15.3 (4637) | 19.3 (1285) | - |
|  | ≥65 | 27.4 (8289) | 28.0 (1862) | - |
| Sex, % (*n*) | |  |  |  |
|  | Male | 50.4 (15257) | 46.2 (3073) | <0.001 |
|  | Female | 49.6 (15032) | 53.8 (3580) | - |
| Social grade, % (*n*) | |  |  |  |
|  | ABC1 | 61.4 (18601) | 60.7 (4036) | 0.110 |
|  | C2DE | 38.6 (11688) | 36.4 (2424) | - |
|  | Missing | 0 (0) | 2.9 (193) |  |
| Region in England, % (*n*) | |  |  |  |
|  | London | 16.8 (5083) | 15.1 (1005) | <0.001 |
|  | South | 22.7 (6885) | 26.1 (1735) | - |
|  | Central | 31.2 (9456) | 29.7 (1974) | - |
|  | North | 29.3 (8865) | 29.1 (1939) | - |
| Before, August 2018 – February 2020. After, April – July 2020. | | | | |

| **Table 2.** Tests of established associations between smoking, high-risk drinking, and sociodemographic characteristics | | | | |
| --- | --- | --- | --- | --- |
|  |  | **%**  **[95% CI]** | **OR**  **[95% CI]** | ***p*** |
| **High-risk drinking** | |  |  |  |
|  | Non-smoker | 34.2 [33.0-35.5] | 1.00 | - |
|  | Smoker | 45.3 [42.1-48.6] | 1.59 [1.38-1.83] | <0.001 |
| **High-risk drinking** | |  |  |  |
|  | Women | 27.4 [26.0-28.9] | 1.00 | - |
|  | Men | 45.5 [43.7-47.2] | 2.21 [1.99-2.44] | <0.001 |
| **Smoking** | |  |  |  |
|  | Age ≥35 | 24.9 [22.7-27.2] | 1.00 | - |
|  | Age 18-34 | 11.2 [10.3-12.0] | 2.65 [2.29-3.07] | <0.001 |
| **Smoking** | |  |  |  |
|  | Social grade ABC1 | 10.8 [9.9-11.8] | 1.00 | - |
|  | Social grade C2DE | 20.0 [18.4-21.6] | 2.06 [1.79-2.37] | <0.001 |
