## Supplementary File 3 for "Moderators of changes in smoking, drinking, and quitting behaviour associated with the first Covid-19 lockdown in England"

Tests of moderation by age, sex, and social grade: interaction results

| **Table 1.** Tests of moderation by age, sex, and social grade: smoking outcomes | | | | | | | | | | | | | | |
| --- | --- | --- | --- | --- | --- | --- | --- | --- | --- | --- | --- | --- | --- | --- |
|  |  | **Age x month x year interaction** | | | | | | | | | | | | |
|  |  | 35-59 [ref 18-34] | | | | | |  | | ≥60 [ref 18-24] | | | | |
|  |  | **OR [95% CI]** | ***p*** |  | **OR_adj_ [95% CI]** | ***p*** |  | | **OR [95% CI]** | | ***p*** |  | **OR_adj_ [95% CI]** | ***p*** |
| Smoking prevalence^1^ | | 0.79 [0.61-1.02] | 0.066 |  | 0.79 [0.61-1.02] | 0.067 |  | | 0.60 [0.43-0.83] | | 0.002 |  | 0.60 [0.43-0.83] | 0.002 |
| Cessation^2^ | | 0.59 [0.20-1.77] | 0.345 |  | 0.59 [0.20-1.78] | 0.347 |  | | 0.54 [0.11-2.68] | | 0.451 |  | 0.52 [0.11-2.58] | 0.423 |
| Quit attempts^2^ | | 0.37 [0.23-0.60] | <0.001 |  | 0.37 [0.23-0.61] | <0.001 |  | | 0.39 [0.20-0.77] | | 0.007 |  | 0.38 [0.19-0.76] | 0.006 |
| Quit success^3^ | | 0.93 [0.28-3.09] | 0.909 |  | 0.96 [0.29-3.22] | 0.946 |  | | 1.08 [0.19-6.26] | | 0.932 |  | 0.89 [0.15-5.23] | 0.893 |
| Use of evidence-based support^3a^ | | 0.48 [0.21-1.12] | 0.090 |  | 0.61 [0.26-1.47] | 0.274 |  | | 0.31 [0.09-1.04] | | 0.058 |  | 0.33 [0.09-1.16] | 0.083 |
| Use of remote support^3b^ | | 0.04 [0.00-1.53] | 0.083 |  | 0.05 [0.00-1.95] | 0.108 |  | | -^4c^ | | - |  | -^4c^ | - |
|  |  | **Sex x month x year interaction** | | | | |  | | **Social grade x month x year interaction** | | | | | |
|  |  | Female [ref male] | | | | |  | | C2DE [ref ABC1] | | | | | |
|  |  | **OR [95% CI]** | ***p*** |  | **OR_adj_ [95% CI]** | ***p*** |  | | **OR [95% CI]** | | ***p*** |  | **OR_adj_ [95% CI]** | ***p*** |
| Smoking prevalence^1^ | | 0.90 [0.72-1.14] | 0.393 |  | 0.91 [0.72-1.14] | 0.395 |  | | 0.85 [0.67-1.07] | | 0.168 |  | 0.85 [0.67-1.07] | 0.171 |
| Cessation^2^ | | 1.52 [0.56-4.17] | 0.413 |  | 1.43 [0.52-3.94] | 0.490 |  | | 1.10 [0.40-3.01] | | 0.858 |  | 1.08 [0.39-2.96] | 0.888 |
| Quit attempts^2^ | | 1.00 [0.64-1.57] | 0.987 |  | 1.01 [0.64-1.58] | 0.969 |  | | 1.20 [0.76-1.88] | | 0.443 |  | 1.21 [0.76-1.91] | 0.422 |
| Quit success^3^ | | 2.04 [0.68-6.10] | 0.204 |  | 2.06 [0.68-6.26] | 0.203 |  | | 0.80 [0.27-2.41] | | 0.697 |  | 0.84 [0.28-2.54] | 0.756 |
| Use of evidence-based support^3a^ | | 1.11 [0.51-2.39] | 0.798 |  | 1.39 [0.62-3.11] | 0.430 |  | | 0.71 [0.32-1.55] | | 0.385 |  | 0.85 [0.38-1.93] | 0.702 |
| Use of remote support^3b^ | | 1.58 [0.10-25.15] | 0.747 |  | 1.64 [0.10-26.69] | 0.730 |  | | -^4d^ | | - |  | -^4d^ | - |
| Note: All data are weighted to match the adult population in England on age, social grade, region, tenure, ethnicity, and working status within sex.  CI, confidence interval. OR, odds ratio. OR_adj_, adjusted odds ratio. The OR_adj_ for smoking prevalence adjusted for trend within year (i.e. August=1 through July=12) and trend across years (i.e. August 2018=1 through July 2020=24). OR_adj_s for other outcomes are additionally adjusted for age, sex, social grade, region (and, for analyses of cessation, quit success, and use of support, heaviness of smoking index).  ^1^ Among all adults.  ^2^ Among past-year smokers.  ^3^ Among past-year smokers who made a quit attempt.  ^a^ Prescription medication, face-to-face behavioural support, nicotine replacement therapy obtained over the counter, e-cigarettes.  ^b^ Telephone support, websites, or apps. ^4^ Estimates could not be produced due to insufficient sample size: just 55 participants reported using remote support in a quit attempt (*n*=14 in the comparator year [*n*=5 Aug-Feb, *n*=9 Apr-Jul]; *n*=41 in the pandemic year [*n*=13 Aug-Feb, *n*=28 Apr-Jul]). ^c^ Of whom just 5 were aged ≥60 years (*n*=1 in the comparator year [*n*=0 Aug-Feb, *n*=1 Apr-Jul]; *n*=4 in the pandemic year [*n*=3 Aug-Feb, *n*=1 Apr-Jul]). ^d^ Of whom 16 were from social grades ABC1 (*n*=2 in the comparator year [*n*=0 Aug-Feb, *n*=2 Apr-Jul]; *n*=14 in the pandemic year [*n*=5 Aug-Feb, *n*=9 Apr-Jul]) and 40 were from C2DE (*n*=12 in the comparator year [*n*=5 Aug-Feb, *n*=7 Apr-Jul]; *n*=28 in the pandemic year [*n*=9 Aug-Feb, *n*=19 Apr-Jul]). | | | | | | | | | | | | | | |

| **Table 2.** Tests of moderation by age, sex, and social grade: drinking outcomes | | | | | | | | | | | | |
| --- | --- | --- | --- | --- | --- | --- | --- | --- | --- | --- | --- | --- |
|  |  | **Age x month x year interaction** | | | | | | | | | | |
|  |  | 35-59 [ref 18-34] | | | | |  | ≥60 [ref 18-24] | | | | |
|  |  | **OR [95% CI]** | ***p*** |  | **OR_adj_ [95% CI]** | ***p*** |  | **OR [95% CI]** | ***p*** |  | **OR_adj_ [95% CI]** | ***p*** |
| High-risk drinking prevalence^1^ | | 1.06 [0.85-1.33] | 0.590 |  | 1.06 [0.85-1.33] | 0.582 |  | 0.98 [0.76-1.27] | 0.880 |  | 0.98 [0.76-1.27] | 0.883 |
| Alcohol reduction attempts^2^ | | 0.56 [0.34-0.92] | 0.022 |  | 0.55 [0.33-0.91] | 0.020 |  | 1.00 [0.53-1.89] | 0.993 |  | 0.98 [0.51-1.86] | 0.942 |
| Use of evidence-based support^3a^ | | 0.34 [0.02-6.21] | 0.469 |  | 0.58 [0.02-13.78] | 0.733 |  | -^4c^ | - |  | -^4c^ | - |
| Use of remote support^3b^ | | 1.58 [0.16-15.82] | 0.697 |  | 1.50 [0.14-16.40] | 0.738 |  | 7.47 [0.23-244.27] | 0.258 |  | 5.06 [0.14-177.22] | 0.372 |
|  |  | **Sex x month x year interaction** | | | | |  | **Social grade x month x year interaction** | | | | |
|  |  | Female [ref male] | | | | |  | C2DE [ref ABC1] | | | | |
|  |  | **OR [95% CI]** | ***p*** |  | **OR_adj_ [95% CI]** | ***p*** |  | **OR [95% CI]** | ***p*** |  | **OR_adj_ [95% CI]** | ***p*** |
| High-risk drinking prevalence^1^ | | 1.32 [1.09-1.61] | 0.005 |  | 1.32 [1.09-1.61] | 0.005 |  | 1.48 [1.21-1.80] | <0.001 |  | 1.48 [1.21-1.80] | <0.001 |
| Alcohol reduction attempts^2^ | | 0.95 [0.60-1.48] | 0.806 |  | 0.98 [0.62-1.54] | 0.932 |  | 0.51 [0.31-0.82] | 0.006 |  | 0.54 [0.33-0.87] | 0.012 |
| Use of evidence-based support^3a^ | | -^4d^ | - |  | -^4d^ | - |  | 0.75 [0.07-8.23] | 0.816 |  | 0.97 [0.07-13.47] | 0.984 |
| Use of remote support^3b^ | | 10.17 [0.84-123.48] | 0.069 |  | 16.88 [1.30-218.96] | 0.031 |  | 0.07 [0.01-0.55] | 0.011 |  | 0.07 [0.01-0.62] | 0.017 |
| Note: All data are weighted to match the adult population in England on age, social grade, region, tenure, ethnicity, and working status within sex.  CI, confidence interval. OR_adj_, adjusted odds ratio. The OR_adj_ for high-risk drinking prevalence is adjusted for trend within year (i.e. August=1 through July=12) and trend across years (i.e. August 2018=1 through July 2020=24). OR_adj_s for other outcomes are additionally adjusted for age, sex, social grade, region (and, for analyses of use of support, full AUDIT score as an indicator of dependence).  ^1^ Among all adults.  ^2^ Among high-risk drinkers.  ^3^ Among high-risk drinkers who made a reduction attempt.  ^a^ Prescription medication or face-to-face behavioural support.  ^b^ Telephone support, websites, or apps. ^4^ Estimates could not be produced due to insufficient sample size: just 54 participants reported using evidence-based support in an alcohol reduction attempt (*n*=34 in the comparator year [*n*=19 Aug-Feb, *n*=15 Apr-Jul]; *n*=20 in the pandemic year [*n*=13 Aug-Feb, *n*=7 Apr-Jul]). ^c^ Of whom just 4 were aged ≥60 years (*n*=4 in the comparator year [*n*=3 Aug-Feb, *n*=1 Apr-Jul]; *n*=4 in the pandemic year [*n*=4 Aug-Feb, *n*=0 Apr-Jul]). ^d^ Of whom 23 were male (*n*=20 in the comparator year [*n*=14 Aug-Feb, *n*=6 Apr-Jul]; *n*=3 in the pandemic year [*n*=3 Aug-Feb, *n*=0 Apr-Jul]) and 31 were female (*n*=14 in the comparator year [*n*=5 Aug-Feb, *n*=9 Apr-Jul]; *n*=17 in the pandemic year [*n*=10 Aug-Feb, *n*=7 Apr-Jul]). | | | | | | | | | | | | |
