## Supplementary File 4 for "Moderators of changes in smoking, drinking, and quitting behaviour associated with the first Covid-19 lockdown in England"

Sensitivity analyses excluding data from July

| **Table 1.** Sample characteristics by survey year and month | | | | | | |
| --- | --- | --- | --- | --- | --- | --- |
|  | | **Comparator year (2018/19)** | |  | **Pandemic year (2019/20)** | |
|  | | August 2018-February 2019 | April-June 2019 |  | August 2019-February 2020 | April-June 2020 |
|  | |  |  |  | (**before lockdown**) | (**during lockdown**) |
| *N* | | 11836 | 4943 |  | 11897 | 4977 |
| Age in years, % (*n*) | |  |  |  |  |  |
|  | 18-24 | 12.9 (1526) | 12.4 (614) |  | 12.7 (1509) | 1009 (545) |
|  | 25-34 | 17.1 (2027) | 17.2 (848) |  | 17.1 (2035) | 17.1 (853) |
|  | 35-44 | 15.8 (1874) | 16.0 (791) |  | 15.8 (1874) | 16.2 (805) |
|  | 45-54 | 17.4 (2054) | 17.2 (851) |  | 17.1 (2038) | 17.2 (854) |
|  | 55-64 | 14.4 (1701) | 14.6 (722) |  | 14.6 (1739) | 15.2 (759) |
|  | ≥65 | 22.4 (2654) | 22.6 (1117) |  | 22.7 (2702) | 23.3 (1161) |
| Sex, % (*n*) | |  |  |  |  |  |
|  | Male | 49.0 (5798) | 49.2 (2431) |  | 49.2 (5856) | 49.4 (2456) |
|  | Female | 51.0 (6038) | 50.8 (2512) |  | 50.8 (6040) | 50.6 (2520) |
| Social grade, % (*n*) | |  |  |  |  |  |
|  | ABC1 | 55.9 (6618) | 55.5 (2743) |  | 55.5 (6602) | 53.4 (2659) |
|  | C2DE | 44.1 (5218) | 44.5 (2200) |  | 44.5 (5295) | 44.4 (2207) |
|  | Missing | 0 (0) | 0 (0) |  | 0 (0) | 2.2 (111) |
| Region in England, % (*n*) | |  |  |  |  |  |
|  | London | 15.6 (1847) | 15.7 (774) |  | 15.5 (1842) | 15.3 (763) |
|  | South | 26.4 (3122) | 26.3 (1298) |  | 26.4 (3146) | 26.6 (1323) |
|  | Central | 30.3 (3588) | 30.2 (1494) |  | 30.1 (3575) | 30.4 (1514) |
|  | North | 27.7 (3280) | 27.9 (1377) |  | 28.0 (3333) | 27.6 (1376) |
| Note: All data are weighted to match the adult population in England on age, social grade, region, tenure, ethnicity, and working status within sex. In some cases, subgroup *n*s do not sum to the total *n* due to rounding. | | | | | | |

| **Table 2.** Tests of moderation by age, sex, and social grade: smoking outcomes | | | | | | | | | | | | | | |
| --- | --- | --- | --- | --- | --- | --- | --- | --- | --- | --- | --- | --- | --- | --- |
|  |  | **Age x month x year interaction** | | | | | | | | | | | | |
|  |  | 35-59 [ref 18-34] | | | | | |  | | 60+ [ref 18-24] | | | | |
|  |  | **OR [95% CI]** | ***p*** |  | **OR_adj_ [95% CI]** | ***p*** |  | | **OR [95% CI]** | | ***p*** |  | **OR_adj_ [95% CI]** | ***p*** |
| Smoking prevalence^1^ | | 0.78 [0.59-1.03] | 0.084 |  | 0.78 [0.59-1.03] | 0.084 |  | | 0.58 [0.41-0.84] | | 0.003 |  | 0.58 [0.41-0.84] | 0.003 |
| Cessation^2^ | | 0.28 [0.07-1.07] | 0.062 |  | 0.28 [0.07-1.08] | 0.065 |  | | 0.25 [0.04-1.56] | | 0.137 |  | 0.24 [0.04-1.52] | 0.131 |
| Quit attempts^2^ | | 0.39 [0.23-0.67] | 0.001 |  | 0.39 [0.23-0.67] | 0.001 |  | | 0.40 [0.19-0.83] | | 0.014 |  | 0.39 [0.19-0.82] | 0.013 |
| Quit success^3^ | | 0.40 [0.10-1.69] | 0.213 |  | 0.39 [0.09-1.67] | 0.203 |  | | 0.48 [0.06-3.60] | | 0.475 |  | 0.37 [0.05-2.81] | 0.334 |
| Use of evidence-based support^3a^ | | 0.83 [0.33-2.11] | 0.695 |  | 1.00 [0.38-2.62] | 0.996 |  | | 0.52 [0.14-1.92] | | 0.324 |  | 0.52 [0.14-2.02] | 0.346 |
| Use of remote support^3b^ | | 0.05 [0.00-1.99] | 0.110 |  | 0.05 [0.00-2.29] | 0.127 |  | | -^4c^ | | - |  | -^4c^ | - |
|  |  | **Sex x month x year interaction** | | | | |  | | **Social grade x month x year interaction** | | | | | |
|  |  | Female [ref male] | | | | |  | | C2DE [ref ABC1] | | | | | |
|  |  | **OR [95% CI]** | ***p*** |  | **OR_adj_ [95% CI]** | ***p*** |  | | **OR [95% CI]** | | ***p*** |  | **OR_adj_ [95% CI]** | ***p*** |
| Smoking prevalence^1^ | | 0.84 [0.65-1.08] | 0.171 |  | 0.84 [0.65-1.08] | 0.172 |  | | 0.86 [0.66-1.12] | | 0.272 |  | 0.86 [0.67-1.12] | 0.275 |
| Cessation^2^ | | 1.93 [0.61-6.10] | 0.265 |  | 1.81 [0.57-5.78] | 0.317 |  | | 1.17 [0.37-3.70] | | 0.789 |  | 1.13 [0.35-3.58] | 0.840 |
| Quit attempts^2^ | | 1.18 [0.72-1.93] | 0.507 |  | 1.19 [0.73-1.96] | 0.485 |  | | 1.18 [0.71-1.94] | | 0.521 |  | 1.15 [0.70-1.91] | 0.578 |
| Quit success^3^ | | 2.20 [0.63-7.66] | 0.214 |  | 2.41 [0.68-8.51] | 0.173 |  | | 0.92 [0.26-3.19] | | 0.890 |  | 0.92 [0.26-3.24] | 0.901 |
| Use of evidence-based support^3a^ | | 1.25 [0.53-2.92] | 0.613 |  | 1.63 [0.67-3.98] | 0.285 |  | | 0.67 [0.28-1.58] | | 0.359 |  | 0.82 [0.33-2.02] | 0.665 |
| Use of remote support^3b^ | | 1.54 [0.09-28.00] | 0.770 |  | 1.66 [0.09-30.88] | 0.734 |  | | -^4d^ | | - |  | -^4d^ | - |
| Note: All data are weighted to match the adult population in England on age, social grade, region, tenure, ethnicity, and working status within sex.  CI, confidence interval. OR, odds ratio. OR_adj_, adjusted odds ratio. The OR_adj_ for smoking prevalence adjusted for trend within year (i.e. August=1 through June=11) and trend across years (i.e. August 2018=1 through June 2020=23). OR_adj_s for other outcomes are additionally adjusted for age, sex, social grade, region (and, for analyses of cessation, quit success, and use of support, heaviness of smoking index).  ^1^ Among all adults.  ^2^ Among past-year smokers.  ^3^ Among past-year smokers who made a quit attempt.  ^a^ Prescription medication, face-to-face behavioural support, nicotine replacement therapy obtained over the counter, e-cigarettes.  ^b^ Telephone support, websites, or apps. ^4^ Estimates could not be produced due to insufficient sample size: just 49 participants reported using remote support in a quit attempt (*n*=11 in the comparator year [*n*=5 Aug-Feb, *n*=6 Apr-Jun]; *n*=38 in the pandemic year [*n*=13 Aug-Feb, *n*=25 Apr-Jun]). ^c^ Of whom just 4 were aged ≥60 years (*n*=0 in the comparator year; *n*=4 in the pandemic year [*n*=3 Aug-Feb, *n*=1 Apr-Jun]). ^d^ Of whom 14 were from social grades ABC1 (*n*=2 in the comparator year [*n*=0 Aug-Feb, *n*=2 Apr-Jul]; *n*=12 in the pandemic year [*n*=5 Aug-Feb, *n*=7 Apr-Jul]) and 36 were from C2DE (*n*=9 in the comparator year [*n*=5 Aug-Feb, *n*=4 Apr-Jul]; *n*=27 in the pandemic year [*n*=9 Aug-Feb, *n*=18 Apr-Jul]). | | | | | | | | | | | | | | |

| **Table 3.** Tests of moderation by age, sex, and social grade: drinking outcomes | | | | | | | | | | | | |
| --- | --- | --- | --- | --- | --- | --- | --- | --- | --- | --- | --- | --- |
|  |  | **Age x month x year interaction** | | | | | | | | | | |
|  |  | 35-59 [ref 18-34] | | | | |  | 60+ [ref 18-24] | | | | |
|  |  | **OR [95% CI]** | ***p*** |  | **OR_adj_ [95% CI]** | ***p*** |  | **OR [95% CI]** | ***p*** |  | **OR_adj_ [95% CI]** | ***p*** |
| High-risk drinking prevalence^1^ | | 1.03 [0.81-1.31] | 0.830 |  | 1.03 [0.81-1.31] | 0.825 |  | 0.98 [0.74-1.31] | 0.909 |  | 0.98 [0.74-1.31] | 0.911 |
| Alcohol reduction attempts^2^ | | 0.51 [0.29-0.87] | 0.014 |  | 0.51 [0.29-0.88] | 0.016 |  | 0.96 [0.48-1.93] | 0.906 |  | 0.96 [0.47-1.94] | 0.898 |
| Use of evidence-based support^3a^ | | 0.60 [0.03-11.49] | 0.734 |  | 0.95 [0.04-24.80] | 0.973 |  | -^4c^ | - |  | -^4c^ | - |
| Use of remote support^3b^ | | 2.91 [0.27-31.14] | 0.377 |  | 3.18 [0.27-37.98] | 0.361 |  | 9.31 [0.26-333.78] | 0.222 |  | 6.61 [0.17-260.67] | 0.314 |
|  |  | **Sex x month x year interaction** | | | | |  | **Social grade x month x year interaction** | | | | |
|  |  | Female [ref male] | | | | |  | C2DE [ref ABC1] | | | | |
|  |  | **OR [95% CI]** | ***p*** |  | **OR_adj_ [95% CI]** | ***p*** |  | **OR [95% CI]** | ***p*** |  | **OR_adj_ [95% CI]** | ***p*** |
| High-risk drinking prevalence^1^ | | 1.38 [1.11-1.71] | 0.004 |  | 1.38 [1.11-1.71] | 0.004 |  | 1.52 [1.22-1.89] | <0.001 |  | 1.52 [1.22-1.89] | <0.001 |
| Alcohol reduction attempts^2^ | | 0.89 [0.55-1.45] | 0.647 |  | 0.92 [0.56-1.50] | 0.733 |  | 0.51 [0.30-0.86] | 0.012 |  | 0.53 [0.31-0.90] | 0.019 |
| Use of evidence-based support^3a^ | | -^4d^ | - |  | -^4d^ | - |  | 1.34 [0.11-16.42] | 0.821 |  | 2.04 [0.13-32.78] | 0.614 |
| Use of remote support^3b^ | | 9.43 [0.75-119.15] | 0.083 |  | 15.85 [1.15-218.64] | 0.039 |  | 0.04 [0.00-0.32] | 0.003 |  | 0.03 [0.00-0.35] | 0.005 |
| Note: All data are weighted to match the adult population in England on age, social grade, region, tenure, ethnicity, and working status within sex.  CI, confidence interval. OR_adj_, adjusted odds ratio. The OR_adj_ for high-risk drinking prevalence is adjusted for trend within year (i.e. August=1 through July=12) and trend across years (i.e. August 2018=1 through July 2020=24). OR_adj_s for other outcomes are additionally adjusted for age, sex, social grade, region (and, for analyses of use of support, full AUDIT score as an indicator of dependence).  ^1^ Among all adults.  ^2^ Among high-risk drinkers.  ^3^ Among high-risk drinkers who made a reduction attempt.  ^a^ Prescription medication or face-to-face behavioural support.  ^b^ Telephone support, websites, or apps. ^4^ Estimates could not be produced due to insufficient sample size: just 50 participants reported using evidence-based support in an alcohol reduction attempt (*n*=31 in the comparator year [*n*=19 Aug-Feb, *n*=12 Apr-Jul]; *n*=19 in the pandemic year [*n*=13 Aug-Feb, *n*=6 Apr-Jul]). ^c^ Of whom just 4 were aged ≥60 years (*n*=3 in the comparator year [*n*=3 Aug-Feb, *n*=0 Apr-Jul]; *n*=4 in the pandemic year [*n*=4 Aug-Feb, *n*=0 Apr-Jul]). ^d^ Of whom 21 were male (*n*=18 in the comparator year [*n*=14 Aug-Feb, *n*=4 Apr-Jul]; *n*=3 in the pandemic year [*n*=3 Aug-Feb, *n*=0 Apr-Jul]) and 29 were female (*n*=13 in the comparator year [*n*=5 Aug-Feb, *n*=8 Apr-Jul]; *n*=16 in the pandemic year [*n*=10 Aug-Feb, *n*=6 Apr-Jul]). | | | | | | | | | | | | |

| **Table 4.** Month (August-February vs. April-June) x year (2018/19 vs. 2019/20) interactions for smoking outcomes | | | | | | | | | | | | | |
| --- | --- | --- | --- | --- | --- | --- | --- | --- | --- | --- | --- | --- | --- |
|  |  | **Prevalence: % [95% CI]** | | | | |  | | | **Month x year interaction** | | | |
|  |  | **2018/19** | |  | **2019/20** | | |  | |  |  |  |  |
|  |  | **Aug-Feb** | **Apr-Jun** |  | **Aug-Feb** | **Apr-Jun** | | |  | **OR [95% CI]** | ***p*** | **OR_adj_**  **[95% CI]** | ***p*** |
| Smoking prevalence^1^ | | 16.9  [16.2-17.5] | 16.3  [15.3-17.4] |  | 16.1  [15.4-16.7] | 16.8  [15.7-17.8] | | |  | 1.09  [0.96-1.24] | 0.168 | 1.09  [0.96-1.24] | 0.164 |
|  | Age 18-34 | 22.4  [21.1-23.8] | 22.6  [20.5-24.8] |  | 21.5  [20.2-22.9] | 27.5  [25.2-29.9] | | |  | 1.37  [1.12-1.68] | 0.002 | 1.37  [1.12-1.68] | 0.002 |
|  | Age 35-59 | 17.5  [16.4-18.6] | 16.0  [14.4-17.6] |  | 16.4  [15.3-17.4] | 15.7  [14.1-17.3] | | |  | 1.06  [0.87-1.30] | 0.555 | 1.06  [0.87-1.30] | 0.549 |
|  | Age ≥60 | 9.9  [8.9-10.9] | 10.2  [8.6-11.8] |  | 10.0  [9.0-11.0] | 8.3  [6.9-9.8] | | |  | 0.79  [0.59-1.07] | 0.131 | 0.80  [0.59-1.07] | 0.134 |
| Cessation^2^ | | 4.0  [3.2-4.9] | 2.9  [1.7-4.0] |  | 3.9  [3.0-4.7] | 9.8  [7.7-11.8] | | |  | 3.79  [2.14-6.72] | <0.001 | 3.70  [2.08-6.58] | <0.001 |
| Quit attempts^2^ | | 28.8  [26.9-30.8] | 27.3  [24.2-30.4] |  | 29.5  [27.5-31.5] | 37.7  [34.6-40.9] | | |  | 1.57  [1.23-2.00] | <0.001 | 1.52  [1.19-1.94] | 0.001 |
|  | Age 18-34 | 32.4  [29.2-35.6] | 24.3  [19.6-28.9] |  | 32.1  [28.9-35.4] | 44.7  [40.0-49.4] | | |  | 2.55  [1.75-3.72] | <0.001 | 2.58  [1.76-3.76] | <0.001 |
|  | Age 35-59 | 27.7  [24.7-30.7] | 31.4  [26.4-36.4] |  | 28.6  [25.5-31.8] | 32.8  [27.8-37.7] | | |  | 1.01  [0.69-1.49] | 0.942 | 1.00  [0.68-1.48] | 0.993 |
|  | Age ≥60 | 22.7  [18.2-27.3] | 25.4  [18.2-32.7] |  | 25.9  [21.2-30.6] | 29.6  [22.0-37.3] | | |  | 1.04  [0.55-1.96] | 0.903 | 1.01  [0.53-1.93] | 0.968 |
| Quit success^3^ | | 12.5  [9.7-15.2] | 12.5  [9.7-15.2] |  | 12.7  [9.9-15.4] | 24.3  [19.5-29.0] | | |  | 2.60  [1.40-4.82] | 0.003 | 2.64  [1.41-4.95] | 0.003 |
| Use of evidence-based support^3a^ | | 53.1  [49.0-57.2] | 50.9  [44.0-57.7] |  | 53.8  [49.7-58.0] | 44.6  [39.1-50.1] | | |  | 0.75  [0.49-1.15] | 0.190 | 0.89  [0.57-1.39] | 0.603 |
| *Table continued on next page.* | | | | | | | | | | | | | |

| **Table 4.** *(continued)* | | | | | | | | | | |
| --- | --- | --- | --- | --- | --- | --- | --- | --- | --- | --- |
| Use of remote support^3b^ | 0.8  [0.1-1.5] | 3.0  [0.7-5.4] |  | 2.4  [1.2-3.7] | 7.9  [5.0-10.9] |  | 0.88  [0.22-3.55] | 0.859 | 0.84  [0.20-3.45] | 0.809 |
| Note: All data are weighted to match the adult population in England on age, social grade, region, tenure, ethnicity, and working status within sex.  CI, confidence interval. OR, odds ratio. OR_adj_, adjusted odds ratio. The OR_adj_ for smoking prevalence adjusted for trend within year (i.e. August=1 through July=12) and trend across years (i.e. August 2018=1 through June 2020=23). OR_adj_s for other outcomes are additionally adjusted for age, sex, social grade, region (and, for analyses of cessation, quit success, and use of support, heaviness of smoking index).  ^1^ Among all adults (2018/19: Aug-Feb *n*=11,820, April-June *n*=4,942; 2019/20: Aug-Feb *n*=11,892, April-June *n*=4,958).  ^2^ Among past-year smokers (2018/19: Aug-Feb *n*=2,055, April-June *n*=818; 2019/20: Aug-Feb *n*=1,961, April-June *n*=925).  ^3^ Among past-year smokers who made a quit attempt (2018/19: Aug-Feb *n*=571, April-June *n*=207; 2019/20: Aug-Feb *n*=551, April-June *n*=317).  ^a^ Prescription medication, face-to-face behavioural support, nicotine replacement therapy obtained over the counter, e-cigarettes.  ^b^ Telephone support, websites, or apps. | | | | | | | | | | |

| **Table 5.** Month (August-February vs. April-June) x year (2018/19 vs. 2019/20) interactions for drinking outcomes | | | | | | | | | | | |
| --- | --- | --- | --- | --- | --- | --- | --- | --- | --- | --- | --- |
|  |  | **Prevalence: % [95% CI]** | | | | |  | **Month x year interaction** | | | |
|  |  | **2018/19** | |  | **2019/20** | |  |  |  |  |  |
|  |  | **Aug-Feb** | **Apr-Jun** |  | **Aug-Feb** | **Apr-Jun** |  | **OR [95% CI]** | ***p*** | **OR_adj_**  **[95% CI]** | ***p*** |
| High-risk drinking prevalence^1^ | | 26.8  [26.0-27.6] | 24.5  [23.3-25.7] |  | 25.8  [25.0-26.6] | 36.9  [35.6-38.3] |  | 1.90  [1.72-2.11] | <0.001 | 1.91  [1.72-2.12] | <0.001 |
|  | Male | 34.6  [33.4-35.8] | 33.3  [31.4-35.1] |  | 34.2  [33.0-35.4] | 45.6  [43.6-47.6] |  | 1.71  [1.49-1.97] | <0.001 | 1.71  [1.49-1.97] | <0.001 |
|  | Female | 19.4  [18.4-20.4] | 16.0  [14.5-17.4] |  | 17.7  [16.7-18.7] | 28.5  [26.8-30.3] |  | 2.36  [2.00-2.78] | <0.001 | 2.36  [2.00-2.78] | <0.001 |
|  | Social grade ABC1 | 31.5  [30.4-32.6] | 30.1  [28.4-31.9] |  | 31.2  [30.1-32.4] | 41.5  [39.6-43.3] |  | 1.66  [1.45-1.90] | <0.001 | 1.66  [1.45-1.90] | <0.001 |
|  | Social grade C2DE | 20.9  [19.8-22.0] | 17.4  [15.8-19.0] |  | 19.1  [18.0-20.1] | 32.1  [30.1-34.1] |  | 2.53  [2.13-3.00] | <0.001 | 2.53  [2.13-3.01] | <0.001 |
| Alcohol reduction attempts^2^ | | 14.8  [13.5-16.0] | 16.1  [14.0-18.2] |  | 15.1  [13.8-16.4] | 26.8  [24.7-28.9] |  | 1.86  [1.47-2.35] | <0.001 | 1.96  [1.54-2.49] | <0.001 |
|  | Age 18-34 | 12.5  [10.6-14.4] | 12.9  [9.6-16.2] |  | 11.7  [9.8-13.7] | 26.8  [23.0-30.6] |  | 2.65  [1.72-4.08] | <0.001 | 2.67  [1.72-4.12] | <0.001 |
|  | Age 35-59 | 16.0  [14.0-17.9] | 19.4  [16.0-22.7] |  | 18.8  [16.7-20.9] | 28.1  [25.0-31.1] |  | 1.34  [0.96-1.86] | 0.085 | 1.39  [0.99-1.94] | 0.055 |
|  | Age ≥60 | 16.5  [13.5-19.5] | 14.0  [9.4-18.6] |  | 13.1  [10.4-15.8] | 24.0  [19.6-28.3] |  | 2.54  [1.46-4.42] | 0.001 | 2.50  [1.43-4.38] | 0.001 |
|  | Social grade ABC1 | 17.4  [15.8-19.1] | 16.2  [13.7-18.8] |  | 16.8  [15.2-18.5] | 30.2  [27.4-32.9] |  | 2.32  [1.75-3.08] | <0.001 | 2.30  [1.73-3.05] | <0.001 |
|  | Social grade C2DE | 9.7  [7.9-11.5] | 15.7  [11.9-19.5] |  | 11.7  [9.6-13.7] | 21.4  [18.3-24.6] |  | 1.19  [0.77-1.85] | 0.441 | 1.25  [0.80-1.96] | 0.327 |
| Use of evidence-based support^3a^ | | 4.3  [2.4-6.1] | 6.3  [2.8-9.8] |  | 3.0  [1.4-4.6] | 1.4  [0.3-2.4] |  | 0.30  [0.09-1.01] | 0.052 | 0.41  [0.10-1.63] | 0.205 |
| *Table continued on next page.* | | | | | | | | | | | |

| **Table 5.** *(continued)* | | | | | | | | | | | |
| --- | --- | --- | --- | --- | --- | --- | --- | --- | --- | --- | --- |
|  | | 3.8  [2.1-5.6] | 6.9  [3.3-10.6] |  | 5.9  [3.7-8.1] | 7.4  [5.0-9.8] |  | 0.68  [0.27-1.68] | 0.399 | 1.08  [0.93-1.27] | 0.308 |
|  | Male | 4.1  [1.8-6.4] | 10.1  [4.6-15.6] |  | 5.3  [2.6-7.9] | 5.3  [2.6-8.1] |  | 0.39  [0.13-1.19] | 0.097 | 0.38  [0.12-1.25] | 0.111 |
|  | Female | 3.4  [0.7-6.2] | 1.5  [0.0-4.4] |  | 6.9  [3.1-10.8] | 10.2  [5.8-14.5] |  | 3.63  [0.37-35.18] | 0.267 | 8.37  [0.75-93.35] | 0.084 |
|  | Social grade ABC1 | 4.3  [2.2-6.4] | 4.1  [0.6-7.5] |  | 4.9  [2.6-7.2] | 8.6  [5.5-11.7 |  | 1.95  [0.59-6.44] | 0.272 | 2.24  [0.64-7.77] | 0.206 |
|  | Social grade C2DE | 2.2  [0.0-5.1] | 13.6  [4.4-22.9] |  | 8.9  [3.6-14.2] | 4.6  [1.1-8.2] |  | 0.07  [0.01-0.44] | 0.005 | 0.07  [0.01-0.57] | 0.014 |
| Note: All data are weighted to match the adult population in England on age, social grade, region, tenure, ethnicity, and working status within sex.  CI, confidence interval. OR, odds ratio. OR_adj_, odds ratio. The OR_adj_ for high-risk drinking prevalence is adjusted for trend within year (i.e. August=1 through June=11) and trend across years (i.e. August 2018=1 through June 2020=23). OR_adj_s for other outcomes are additionally adjusted for age, sex, social grade, region (and, for analyses of use of support, full AUDIT score as an indicator of dependence).  ^1^ Among all adults (2018/19: Aug-Feb *n*=11,793, April-June *n*=4,920; 2019/20: Aug-Feb *n*=11,828, April-June *n*=4,884).  ^2^ Among high-risk drinkers (2018/19: Aug-Feb *n*=3,091, April-June *n*=1,159; 2019/20: Aug-Feb *n*=2,986, April-June *n*=1,713).  ^3^ Among high-risk drinkers who made a reduction attempt (2018/19: Aug-Feb *n*=456, April-June *n*=187; 2019/20: Aug-Feb *n*=449, April-June *n*=453).  ^a^ Prescription medication or face-to-face behavioural support.  ^b^ Telephone support, websites, or apps. | | | | | | | | | | | |
